# A minimally Invasive Biomarker for Sensitive and Accurate Diagnosis of Parkinson’s Disease

**DOI:** 10.1101/2024.06.29.24309703

**Authors:** Zerui Wang, Tricia Gilliland, Hyun Jo Kim, Maria Gerasimenko, Kailey Sajewski, Manuel V. Camacho, Gurkan Bebek, Shu G. Chen, Steven A. Gunzler, Qingzhong Kong

## Abstract

**Importance:** Parkinson’s disease (PD), the second most common neurodegenerative disease, is pathologically characterized by intraneuronal deposition of misfolded alpha-synuclein aggregates (αSyn^D^). αSyn^D^ seeding activities in CSF and skin samples have shown great promise in PD diagnosis, but they require invasive procedures. Sensitive and accurate αSyn^D^ seed amplification assay (αSyn-SAA) for more accessible and minimally invasive samples (such as blood and saliva) are urgently needed for PD pathological diagnosis in routine clinical practice.

**Objective:** To develop a sensitive and accurate αSyn-SAA biomarker using blood and saliva samples for sensitive, accurate and minimally invasive PD diagnosis.

**Design, Setting, and Participants:** This prospective diagnostic study evaluates serum and saliva samples collected from patients clinically diagnosed with PD or healthy controls (HC) without PD at an academic Parkinson’s and Movement Disorders Center from February 2020 to March 2024. Patients diagnosed with non-PD parkinsonism were excluded from this analysis. A total of 124 serum samples (82 PD and 42 HC) and 131 saliva samples (83 PD and 48 HC) were collected and examined by αSyn-SAA. Out of the 124 serum donors, a subset of 74 subjects (48 PD and 26 HC) also donated saliva samples during the same visits. PD patients with serum samples had a mean age of 69.21 years (range 44-88); HC subjects with serum samples had a mean age of 66.55 years (range 44-81); PD patients with saliva samples had a mean age of 69.58 years (range 49-87); HC subjects with saliva samples had a mean age of 64.71 years (range 30-81).

**Main Outcomes and Measures:** Serum and/or saliva αSyn^D^ seeding activities from PD and HC subjects were measured by αSyn-SAA using the Real-Time Quaking-Induced Conversion (RT-QuIC) platform. These PD patients had extensive clinical assessments including MDS-UPDRS. For a subset of PD and HC subjects whose serum and saliva samples were both collected during the same visits, the αSyn^D^ seeding activities in both samples from the same subjects were examined, and the diagnostic accuracies for PD based on the seeding activities in either sample alone or both samples together were compared.

**Results:** RT-QuIC analysis of αSyn^D^ seeding activities in the 124 serum samples revealed a sensitivity of 80.49%, a specificity of 90.48%, and an accuracy of 0.9006 (AUC of ROC, 95% CI, 0.8472-0.9539, *p*<0.0001) for PD diagnosis. RT-QuIC analysis of αSyn^D^ seeding activity in 131 saliva samples revealed a sensitivity of 74.70%, a specificity of 97.92%, and an accuracy of 0.8966 (AUC of ROC, 95% CI, 0.8454-0.9478, *p*<0.0001). When aSyn^D^ seeding activities in the paired serum-saliva samples from the subset of 48 PD and 26 HC subjects were considered together, sensitivity was 95.83%, specificity was 96.15%, and the accuracy was 0.98 (AUC of ROC, 95% CI, 0.96-1.00, *p*<0.001), which are significantly better than when αSyn^D^ seeding activities in either serum or saliva were used alone. For the paired serum-saliva samples, when specificity was set at 100% by elevating the αSyn-SAA cutoff values, a sensitivity of 91.7% and an accuracy of 0.9457 were still attained. Detailed correlation analysis revealed that αSyn^D^ seeding activities in the serum of PD patients were correlated inversely with Montreal Cognitive Assessment (MoCA) score (*p*=0.04), positively with Hamilton Depression Rating Scale (HAM-D) (*p*=0.03), and weakly positively with PDQ-39 cognitive impairment score (*p*=0.07). Subgroup analysis revealed that the inverse correlation with MoCA was only seen in males (*p*=0.013) and weakly in the ≥70 age group (*p*=0.07), and that the positive correlation with HAM-D was only seen in females (*p*=0.04) and in the <70 age group (*p*=0.01). In contrast, αSyn^D^ seeding activities in the saliva of PD patients were inversely correlated with age at diagnosis (*p*=0.02) and the REM sleep behavior disorder (RBD) status (*p*=0.04), but subgroup analysis showed that the inverse correlation with age at diagnosis was only seen in males (*p*=0.04) and in the <70 age group (*p*=0.01).

**Conclusion and Relevance:** Our data show that concurrent RT-QuIC assay of αSyn^D^ seeding activities in both serum and saliva can achieve high diagnostic accuracies comparable to that of CSF αSyn-SAA, suggesting that αSyn^D^ seeding activities in serum and saliva together can potentially be used as a valuable biomarker for highly sensitive, accurate, and minimally invasive diagnosis of PD in routine clinical practice. αSyn^D^ seeding activities in serum and saliva of PD patients correlate differentially with some clinical characteristics and in an age and sex-dependent manner.

**KEY POINTS:** *Question:* Are αSyn^D^ seeding activities in serum and saliva together a more sensitive and accurate diagnostic PD biomarker than αSyn^D^ seeding activities in either sample type alone? Are αSyn^D^ seeding activities in either serum or saliva correlated with any clinical characteristics?

*Findings:* Examinations of αSyn^D^ seeding activities in 124 serum samples and 131 saliva samples from PD and heathy control subjects show that αSyn^D^ seeding activities in both serum and saliva samples together can provide significantly more sensitive and accurate diagnosis of PD than either sample type alone. αSyn^D^ seeding activities in serum or saliva exhibit varied inverse or positive correlations with some clinical features in an age and sex-dependent manner.

*Meaning:* αSyn^D^ seeding activities in serum and saliva together can potentially be used as a valuable pathological biomarker for highly sensitive, accurate, and minimally invasive PD diagnosis in routine clinical practice and clinical studies, and αSyn^D^ seeding activities in serum or saliva correlate with some clinical characteristics in an age and sex-dependent manner, suggesting some possible clinical utility of quantitative serum/saliva αSyn-SAA data.

## INTRODUCTION

Parkinson’s disease (PD) is the second most common neurodegenerative disease that affects almost 1 million Americans and >10 million patients globally according to Parkinson’s Foundation. However, accurate diagnosis for PD remains a challenge. The clinical diagnosis of PD is based on clinical examination with a diagnostic accuracy of ∼80% at early stages, dependent on an expert clinician and time-consuming MDS-UPDRS examination (Hughes et al., 2002, Adler et al. 2014, Rizzo et al. 2016, Virameteekul et al. 2023). A definitive diagnosis for PD still relies on postmortem detection of neuronal inclusions of misfolded and aggregated alpha-synuclein protein (αSyn) in autopsy brain tissues.

αSyn plays a critical role in PD pathogenesis, and the deposition of disease-associated α-synuclein aggregates (αSyn^D^) in the central nervous system is the pathological hallmark of PD and other synucleinopathies, such as multiple system atrophy (MSA) and dementia with Lewy bodies (DLB). The recently developed αSyn seed amplification assay (αSyn-SAA) can detect αSyn^D^ at extremely high sensitivity and specificity utilizing the ultrasensitive real-time quaking-induced conversion (RT-QuIC) platform (Groveman et al. 2018, Srivastava et al. 2022, Yoo et al. 2022). The sample types that have been examined by aSyn-SAA assays include CSF (Fairfoul et al. 2016, Kang et al. 2019, Orru et al. 2021, Russo et al. 2021, Bräuer et al. 2023, Concha-Marambio et al. 2023a/b, Oftedal et al. 2023, Siderwowf et al. 2023, Brockmann et al. 2024), olfactory mucosa (Stefani et al. 2021, Bargar at al. 2021b), biopsy or autopsy skin (Manne et al. 2020a, Wang et al. 2020, Bargar et al. 2021a/b; Kuzkina et al. 2021), salivary gland (Manne et al. 2020b, Bargar et al. 2021a), saliva (Luan et al. 2022, Vivacqua et al. 2023), and blood (Kluge et al. 2022 & 2024, Okuzumi et al. 2023, Schaeffer et al. 2024). CSF, skin, and salivary gland αSyn-SAAs studies have shown excellent results for PD diagnosis. CSF αSyn-SAA demonstrated an impressive sensitivity of 87.7% for PD and a specificity of 96.3% for healthy controls with a large number of CSF samples in a recent rigorous three-lab study (Siderowf et al. 2023), largely in line with various studies reporting 86-96% sensitivities for PD detection. Skin αSyn-SAA also showed very impressive diagnostic accuracy for PD in studies from our groups and others (Manne et al. 2020a, Wang et al. 2020, Bargar et al. 2021a/b; Kuzkina et al. 2021). However, the invasive sampling procedures for CSF and skin biopsy are a significant impediment to patient acceptance and routine clinical application.

Collection of blood and saliva samples is minimally or non-invasive and least expensive, making these samples more desirable for clinical diagnosis. A recent αSyn-SAA study with serum samples showed ∼95% sensitivity for PD and DLB and ∼92% specificity for healthy controls (Okuzumi et al. 2023), but when specificity is set at 95%, the sensitivity would drop to ∼85%, which is comparable to the CSF αSyn-SAA (Siderowf et al. 2023). A more recent article reported an extremely high sensitivity (98.8%) in PD detection with serum samples using a modified aSyn-SAA protocol (Schaffer et al. 2024), but the extraordinary performance of such serum aSyn-SAA in PD diagnosis is yet to be verified by other laboratories using samples from diverse patient cohorts. Saliva has also been reported to be a good source for αSyn^D^ seeding activity from PD patients (Luan et al. 2022, Vivacqua et al. 2023), where the saliva aSyn-SAA assay showed 76.0% sensitivity for PD and 94.4% specificity for healthy controls in one report (Luan et al. 2022) and 83.78% sensitivity and 82.61% specificity in another (Vivacqua et al. 2023). None of the blood or saliva αSyn-SAA assays have been vigorously verified.

Here we report αSyn-SAA analysis of 124 serum samples and 131 saliva samples from PD and healthy control subjects and show that using αSyn^D^ seeding activities in both serum and saliva samples together can achieve much higher sensitivity and specificity for PD diagnosis than using either sample type alone.

## METHODS

### Study Oversight

The study was approved by the University Hospitals Cleveland Medical Center Institutional Review Board. All research subjects had the capacity and gave written informed consent, according to the Declaration of Helsinki.

### Clinical assessments

Diagnostic criteria were applied, and neurological examination was conducted by movement disorders clinicians within the Parkinson’s and Movement Disorders Center at University Hospitals Cleveland Medical Center, in the United States. There were two recruitment mechanisms. 124 subjects (82 PD patents and 42 healthy controls) were recruited in cohort 1 within our skin and peripheral biofluid biomarker study between February 2020 and March 2024. Inclusion criteria included age 21-89 years, with PD onset at or above 40 years of age, and Montreal Cognitive Assessment (MoCA) score of at least 10. Exclusion criteria included a blood clotting disorder, treatment with multiple antiplatelets or anticoagulants, allergy to local anesthetic, deep brain stimulation, or another neurodegenerative disorder. PD patients were required to meet UK Brain Bank Criteria for possible or probable PD (Daniel and Lees 1993, Gelb et al. 1999). Additionally, inclusion criteria for the NIH Parkinson Disease Biomarker Program (PDBP), including MDS Clinical Diagnostic Criteria for PD (Postuma et al. 2015), were also applied in this cohort. The Movement Disorder Society-Unified Parkinson’s Disease Rating Scale (MDS-UPDRS) and modified Hoehn & Yahr (mH&Y) were completed in PD subjects. All subjects had demographics, Schwab & England (S&E), MoCA, Hamilton Anxiety Rating Scale (HAM-A), Hamilton Depression Rating Scale (HAM-D), Epworth Sleepiness Scale (ESS), PDQ-39, REM Behavior Disorder (RBD) Questionnaire, vital signs, family history, and full neurological examination. Most cohort 1 PD patients were also assessed at the one-year follow-up visits, and for these patients the clinical diagnosis of PD was based on the assessment at the one-year visit.

56 subjects (34 with PD and 22 HC controls were recruited for saliva (and some for blood too) and data collection from our saliva and blood biomarker study between May 2023 and May 2024, Inclusion criteria included ages 30-95 years, without an upper respiratory infection, and PD subjects with or without DBS must meet UK Brain Bank criteria for (possible or probable). In this study cohort (cohort 2), subjects had demographics, family history, clinician-determined cognitive status, MDS-UPDRS motor score (or Part 3), and a subset of subjects had MoCA.

Subjects with pregnancy, schizophrenia, negative dopamine transporter SPECT, or neuroleptic-induced parkinsonism were excluded from both studies.

### Blood collection and serum sample preparation

Blood was collected by drawing 3.0 ml of whole blood into a clot activator tube. The blood was allowed to clot by leaving it undisturbed at room temperature for approximately 30 minutes and no more than an hour. The clot was removed by centrifuging at 2,000 x g for 10 minutes. The resulting supernatant (serum) was pipetted into two screw-cap polypropylene cryovials while maintained at 2-8°C, then stored in a -80°C freezer. The stored serum samples were transported on dry ice when needed.

### Saliva sample collection and preparation

Saliva (2-6 ml) was collected from patients after at least 60 minutes of fasting (including no chewing gum), 4 hours without tobacco, and 12 hours without alcohol. Specimens were collected by drooling into a funnel within a 2 ml cryovial. If this was not possible, then it was instead obtained by chewing on a sterile cotton swab for a few minutes and then depositing it in a specimen container. Saliva was immediately stored at -80°C in a cryovial and transported on dry ice when needed.

### Purification of recombinant αSyn

The purification of recombinant αSyn was executed according to previously published methodologies (Becker et al.,2018, Volpicelli-Daley et al., 2014). The BL21 E. coli strain, previously transformed with a plasmid expressing human αSyn, was cultured in Terrific Broth Medium until an optical density of 0.9 was achieved at 600 nm. Cells were then induced using the Overnight Express™ Autoinduction System 1 (Sigma-Aldrich). For purification, the cells were lysed in a high-salt buffer [750 mM NaCl, 10 mM Tris, pH 7.6, 1 mM EDTA, and 1 mM phenylmethylsulfonyl fluoride (PMSF)] and then boiled at 100°C for 20 minutes. Following the removal of cell debris, the lysate underwent overnight dialysis in 10 mM Tris, pH 7.6, 50 mM NaCl, and 1 mM EDTA before being loaded onto a Superdex 75 column (GE Healthcare Life Sciences). α-Syn-containing fractions were identified via SDS-PAGE and Coomassie blue staining, followed by another overnight dialysis in 10 mM Tris, pH 7.6, 25 mM NaCl, and 1 mM EDTA. The protein underwent chromatographic separation using a Hi-Trap Q HP anion-exchange column (GE Healthcare Life Sciences) and eluted using a 0 to 1 M NaCl gradient; α-Syn typically elutes around 350 mM NaCl as observed by absorption at 280 nm. SDS-PAGE and Coomassie blue staining were utilized to confirm the purity of α-Syn. Finally, purified α-Syn was dialyzed into 10 mM Tris (pH 7.6) and 50 mM NaCl, divided into aliquots, and stored at -80°C until required.

### Immunoprecipitation-based RT-QuIC for serum and saliva samples

The Immunoprecipitation (IP)-based RT-QuIC is modified from a previous report (Okuzumi et al., 2023). Saliva samples were centrifuged at 7000g at 4°C for 10 minutes before the IP process, and the supernatant was kept. For immunoprecipitation, 150 µL of lysis buffer (1% BSA, 150 mM NaCl, 1% Triton X, 50 mM Tris-HCl, pH 7, 1 mM PMSF) with 2.5 µg MJFR-14 (anti-α-synuclein conformational antibody: Abcam, UK) and 35 µL protein A/G agarose beads (Thermo Fisher Scientific, USA) was incubated overnight at 4 °C, and 150 µL of serum or saliva supernatant was added and rotated at 4°C for 4 hours. The eluted proteins were precipitated using 4-fold acetone at -20°C for 4 hours, then centrifuged at 13000g for 20 minutes. The pellet was resuspended in seed dilution buffer for RT-QuIC (1x N2 supplement in PBS, pH 7.5). The RT-QuIC reaction conditions were based on previous studies (Wang et al., 2020; Bargar et al., 2021; Russo et al., 2021). The reaction mix contains 100 mM phosphate buffer (pH 8.2), 150 mM NaCl, 20 µM ThT, and 7 µM recombinant <u>α</u>Syn protein. In a 96-well plate (Nunc) pre-loaded with six 0.8 mm low-binding glass beads (OPS Diagnostic LLC, USA); 98 µL of the reaction mix was added per well, followed by 2 µL of saliva/serum IP diluents. Plates were sealed with a film (Nalgene Nunc International) and incubated at 40 °C in a BMG FLUOstar Omega plate reader. The incubation included 1 minute of double orbital shaking at 400 rpm and 15 minutes of rest, repeated for the duration of the assay. ThT fluorescence measurements (450 ± 10 nm excitation and 480 ± 10 nm emission; bottom read) were recorded every 45 minutes. Four replicate reactions were conducted per sample dilution, and the average fluorescence values were calculated. If a sample had only one positive well, the sample was considered negative, and the average of the remaining three wells was used for comparative analysis. The ThT fluorescence readings at the endpoint (93.35 hours) were normalized to a percentage of the maximal possible fluorescence reading (260,000) and used to measure the relative αSynD seeding activities in the respective samples.

### Statistical Analysis

All statistical analyses were performed with the R statistical software (version 4.4.0). Descriptive statistics, including means, standard deviations, and *p* values for continuous variables such as age and clinical test scores, were computed using the R package ‘tableone’ (version 0.13.2) via two-sample t-tests. For categorical variables, such as sex, chi-squared tests were applied to determine the *p* values. Paired serum and saliva samples were systematically analyzed across a range of ThT fluorescence cutoff values for each sample type: from 45,000 to 73,000 for saliva and from 35,000 to 69,000 for serum, incremented by 100. Sensitivity, specificity, and accuracy at each cutoff combination was assessed. To provide more robust estimates, 95% confidence intervals and *p* values were calculated using bootstrap resampling with 1,000 iterations, facilitated by the ‘boot’ package (version 1.3-30). The performance of these classifications was illustrated using ROC curves (Hajian-Tilaki et al. 2013), generated via the ‘ROCR’ package (version 1.0-11). The optimal cutoff values for both serum and saliva were identified by plotting the accuracy against various cutoff combinations in a 3-D space, utilizing the ‘plotly’ package (version 4.10.4).

The relationship between aSyn^D^ seeding activities measured by RT-QuIC and various clinical features was evaluated using Pearson’s correlation coefficient, with the resultant correlations visualized through the ‘ggplot2’ package (version 3.5.1). The RT-QuIC data for the paired serum-saliva samples was also examined and binary features (such as hyposmia and RBD) were examined with ANOVA. For age subgroup analyses, 70 years was chosen as the cutoff age because the age distribution was centered around 70, ensuring a sufficient number of cases in each group to maintain statistical power.

## RESULTS

A total of 124 serum samples (82 PD and 42 HC) and 131 saliva samples (83 PD and 48 HC) were examined by RT-QuIC assay (Table 1). The patient group for the PD serum samples had a mean age of 69.21 years (range 44-88) and included 45 males (54.9%); the patient group for the HC serum samples had a mean age of 66.55 years (range 44-81) and included 11 males (26.2%) (Table 1). The patient group for the PD saliva samples had a mean age of 69.58 years (range 49-87) and included 46 males (55.4%), and the subject group for the HC saliva samples had a mean age of 64.71 years (range 30-81) and included 14 males (29.2%) (Table 1).

**Table 1.** Demographics and clinical features of PD and HC subjects.

|  | All cases |  |  |  | ss-subset |  |
| --- | --- | --- | --- | --- | --- | --- |
|  | PD |  | HC |  | PD | HC |
|  | Serum | Saliva | Serum | Saliva | Serum & Saliva | Serum & Saliva |
| Sample, n | 82 | 83 | 42 | 48 | 48 | 26 |
| Age, mean (range), years | 69.21 (44-88) | 69.58 (49-87) | 66.55 (44-81) | 64.71 (30-81) | 68.83 (49-84) | 66.38 (44-81) |
| Male, number (%) | 45 (54.9) | 46 (55.4) | 11 (26.2) | 14 (29.2) | 25 (52.1) | 7 (26.9) |
| Disease duration, (range), years | 5.05 (0-17) | 6.18 (0-31) | NA | NA | 5.04 (0-17) | NA |
| mH&Y, mean (SD) | 2.1 (0.48) | 2.09 (0.61) | NA | NA | 2.04 (0.52) | NA |
| Self-reported hyposmia | 37 (45.1) | 21 (42.9) | 1 (2.6) | 0 (0) | 20 (41.7) | 0 (0) |
| aSyn-SAA +*, Number (%) | 66 (80.5) | 62 (74.7) | 4 (9.5) | 1 (2.1) | 46 (95.8) | 1 (3.8) |

\*Positive by aSyn-SAA

### Detection of αSyn^D^ seeding activity in serum from PD and HC subjects by RT-QuIC Assay

We developed an immunoprecipitation-based aSyn-SAA protocol with RT-QuIC (modified from a protocol reported earlier by Okuzumi et al. 2023) to detect αSyn^D^ seeding activities in serum samples from PD and HC subjects. Representative RT-QuIC assay curves for blinded saliva and serum samples including 10 PD and 10 HC ach are shown in Figure 1, which indicate that the saliva and serum samples of PD patients had an overall higher ThT fluorescence intensity over the time than those of the healthy controls.

**Figure 1.**
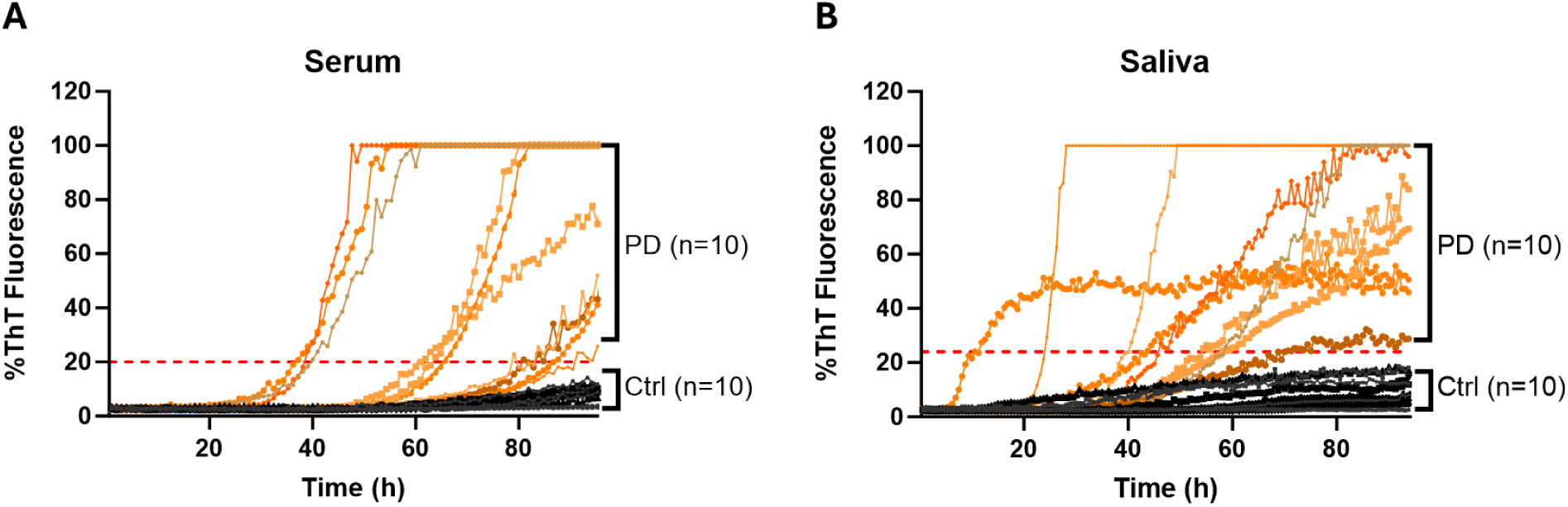
Representative ThT Fluorescence Curves of αSyn RT-QuIC Assays of Serum or Saliva Samples. **A**. Representative curves of ThT fluorescence readings over time for RT-QuIC assays of αSyn^D^ seeding activities in serum samples from 10 PD and 10 HC subjects. **B**. Representative curves of ThT fluorescence readings over time for RT-QuIC assays of αSyn^D^ seeding activities in saliva samples from 10 PD and 10 HC subjects. All samples were coded and blinded for the RT-QuIC assays. The ThT fluorescence readings at the endpoint (93.35 hr) were normalized to percentages of the maximal fluorescence reading (260,000) and used to measure the relative αSyn^D^ seeding activities in the respective samples. Orange lines: curves for PD samples; black lines: curves for HC subjects.

Examination of 124 serum samples from 82 PD patients (63 probable PD and 19 possible PD) and 42 HC subjects revealed a sensitivity of 80.49%, a specificity of 90.48%, and an accuracy of 0.9006 (AUC of ROC, 95% CI, 0.8472-0.9539, *p*<0.0001) for diagnosis of patients with PD compared with clinical diagnosis (Figure 2, Table 2). For serum samples from probable PD patients, the sensitivity, specificity, and accuracy were 79.37%, 90.48%, and 0.8857 (AUC of ROC, 95% CI, 0.8212-9502, *p*<0.0001), respectively (Figure S1-A & B).

**Figure 2.**
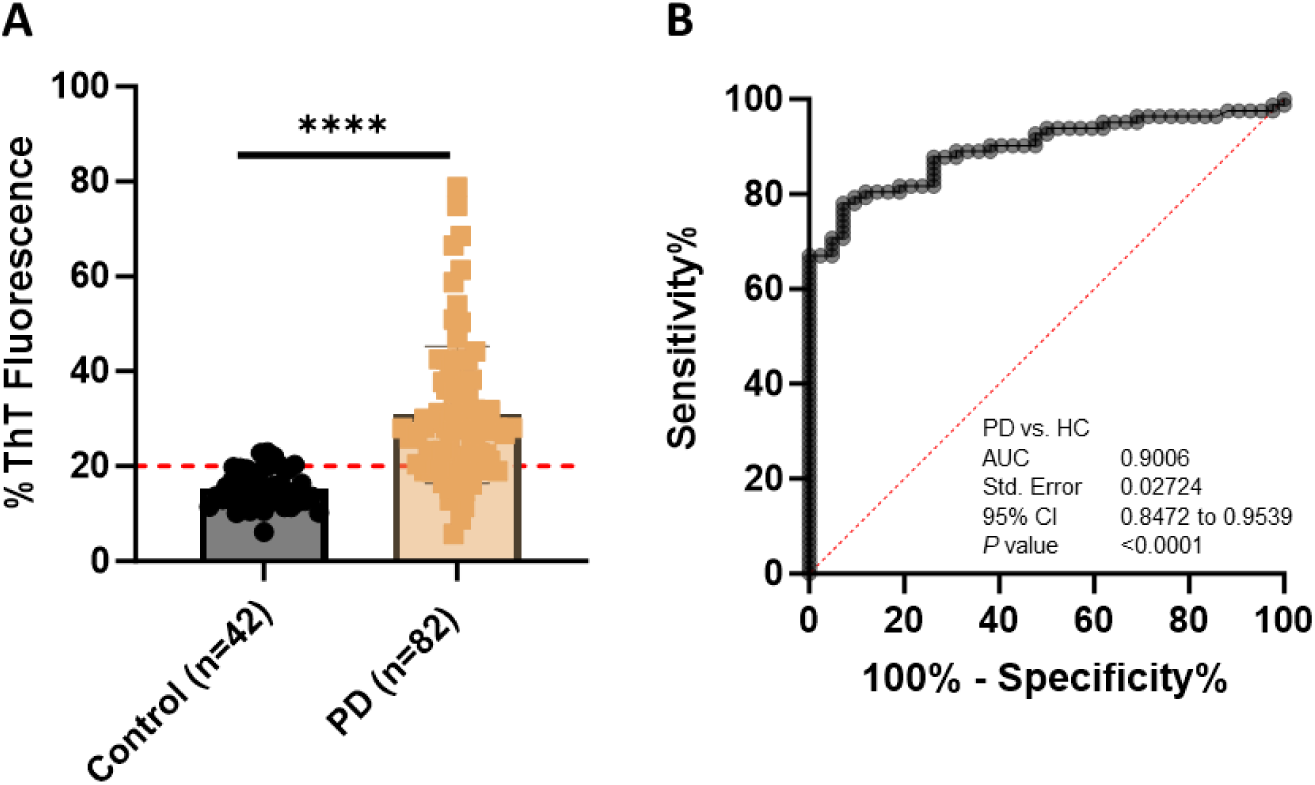
Comparison of αSyn^D^ Seeding Activity in Serum Samples from PD Patients and Healthy Controls (HC) by RT-QuIC. **A**. Scatter graph of RT-QulC ThT fluorescence intensity of αSyn^D^ seeding activity of serum samples from PD patients and HC. Scatter graph of the average of the peak thioflavin T (ThT) fluorescence in quadruplicate wells as a percentage of the maximum fluorescence (%ThT fluorescence) of serum from 82 PD patients and 42 HC subjects by RT-QuIC assay. ThT fluorescence cutoff: 52,105. **** *p*<0.0001. **B**. ROC curve and AUC for αSyn^D^ seeding activity comparisons between PD patients and HC. ROC curve and AUC value were obtained based on αSyn^D^ seeding activity in serum of the PD patients and HC. SE, standard error. 95% CI, 95% confidence interval.

**Table 2.**
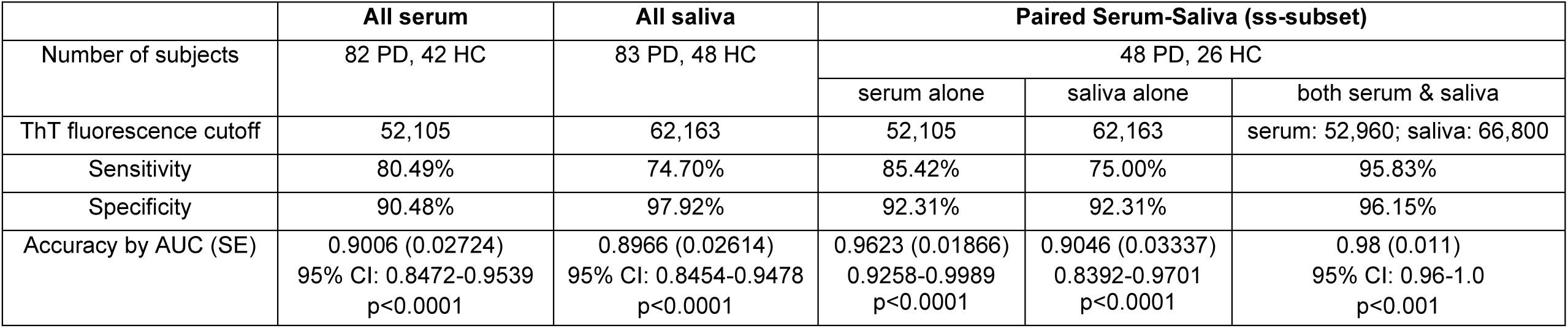
Comparison of diagnostic accuracy for PD with serum αSyn-SAA, saliva αSyn-SAA, or serum αSyn-SAA and saliva αSyn-SAA together.

### Detection of αSyn^D^ seeding activity in saliva from PD and HC subjects by RT-QuIC Assay

We used the same RT-QuIC protocol to examine αSyn^D^ seeding activities in saliva samples from 83 PD (24 probable PD and 59 possible PD) and 48 HC subjects and achieved a sensitivity of 74.70%, a specificity of 97.92%, and an accuracy of 0.8966 (AUC of ROC, 95% CI, 0.8454-0.9478, *p*<0.0001) for diagnosis of all patients with PD (Figure 3, Table 2). For saliva samples from probable PD cases, the sensitivity, specificity, and accuracy were 79.66%, 97.92%, and 0.9054 (AUC of ROC, 95% CI, 0.8484-09623, *p*<0.0001), respectively (Figure S1-C & D)

**Figure 3.**
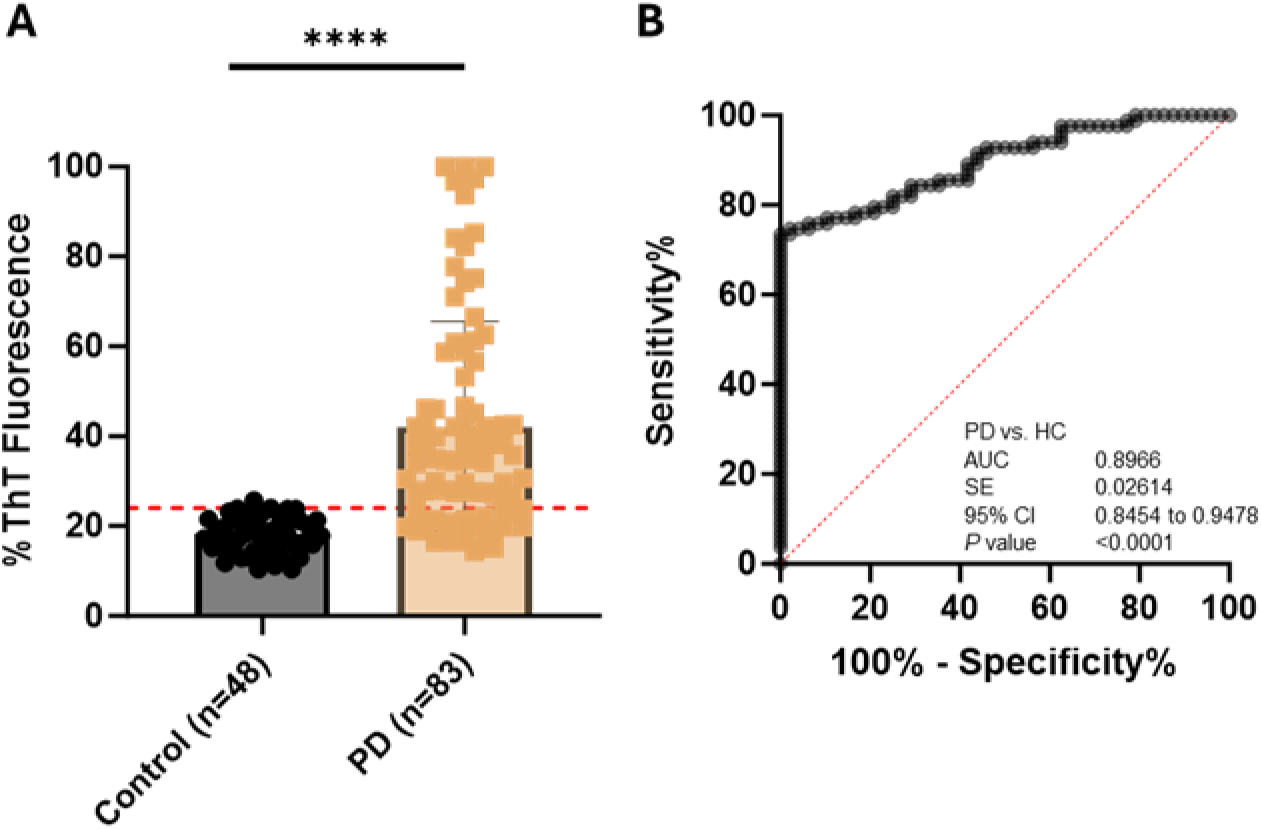
Comparison of α-Syn^D^ Seeding Activity in Saliva Samples from PD Patients and Healthy Control (HC) by RT-QuIC. **A**. Scatter graph of RT-QulC ThT fluorescence intensity of αSyn^D^ seeding activity of serum samples from PD patients and HC. Scatter graph of the average of the peak thioflavin T (ThT) fluorescence in quadruplicate wells as a percentage of the maximum fluorescence (%ThT fluorescence) of saliva from 49 PD patients and 26 HC by RT-QuIC assay. ThT fluorescence cutoff: 62,613. **** *P*<0.0001. **B**. ROC curve and AUC for αSyn^D^ seeding activity comparisons between the PD patients and HC. ROC curve and AUC value for αSyn^D^ seeding activity in saliva of the PD patients and HC. SE, standard error. 95% CI, 95% confidence interval.

### PD diagnosis based on αSyn^D^ seeding activities in a subset of PD and HC subjects with both serum and saliva samples

We hypothesized that PD patients who are αSyn-SAA negative for their serum samples may be αSyn-SAA positive for their saliva samples and vice versa. To test this hypothesis, we conducted αSyn-SAA assays on both serum and saliva samples from a subset (termed “ss-subset”) of 48 PD patients (34 probable PD and 14 possible PD) and 26 HC subjects who provided both blood and saliva samples during the same visits. When either sample type was analyzed separately, a sensitivity of 85.42%, a specificity of 92.31%, and an accuracy of 0.9623 (AUC of ROC, 95% CI, 0.9258-0.9989, *p*<0.0001) were achieved for diagnosis of all PD patients in the ss-subset using serum aSyn-SAA data alone (Figure 4A-B). For serum samples from probable PD cases of the ss-subset, the sensitivity, specificity, and accuracy were 85.29%, 92.31%, and 0.9615 (AUC of ROC, 95% CI, 0.9210-1.000, *p*<0.0001), respectively (Figure S2-A & B). In comparison, a sensitivity of 75.00%, a specificity of 92.31%, and an accuracy of 0.9046 (AUC of ROC, 95% CI, 0.8392-0.9701, *p*<0.0001) were achieved for the diagnosis of all PD patients of the ss-subset using saliva samples alone (Figure 4-C & D, Table 2). For saliva samples from probable PD cases of the ss-subset, the sensitivity, specificity, and accuracy were 76.47%, 92.31%, and 0.8824 (AUC of ROC, 95% CI, 0.7971-09676, *p*<0.0001), respectively (Figure S2-C & D).

**Figure 4.**
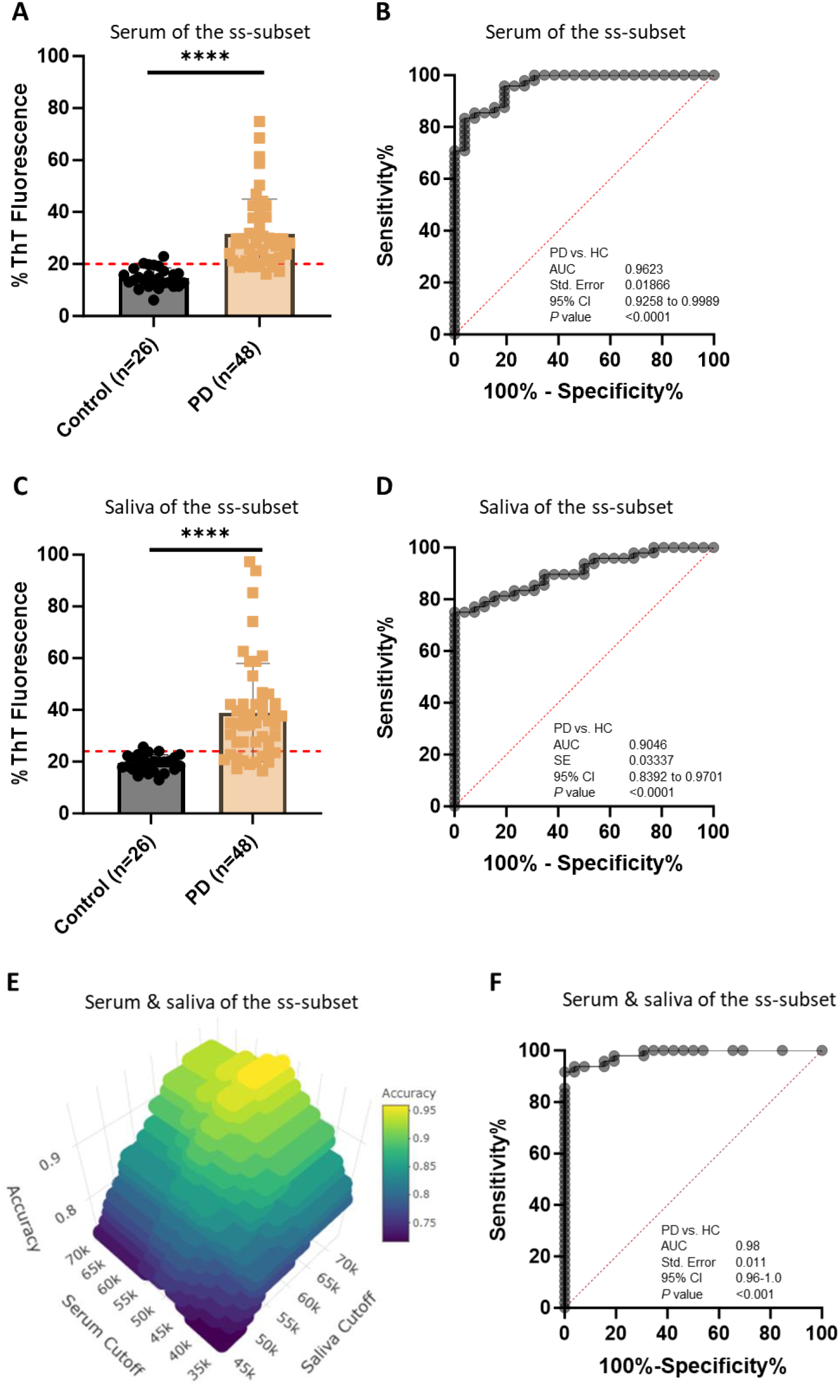
Enhanced Diagnostic Accuracy for PD Using αSyn^D^ Seeding Activities in Both Serum and Saliva Samples from a Subset of PD Patients and Healthy Control (HC) by RT-QuIC. **A**. Scatter graph of αSyn^D^ seeding activities (RT-QuIC ThT fluorescence intensity of serum samples) from in a subset of PD and HC subjects with paired serum and saliva samples. Scatter graph was plotted based on the average of the peak thioflavin T (ThT) fluorescence in quadruplicate wells as a percentage of the maximum fluorescence (%ThT fluorescence) in RT-QuIC assay of serum samples from 48 PD patients and 26 HC in a subset of PD and HC subjects with both serum and saliva samples (termed serum-saliva subset or ss-subset). ThT fluorescence cutoff: 52,105. **** *p*<0.0001. **B**. ROC curve and AUC for serum αSyn^D^ seeding activity comparisons between the PD patients and HC subjects in the ss-subset. ROC curve and AUC value were obtained based on αSyn^D^ seeding activity in serum samples from the PD patients and HC of the ss-subset shown in panel A. **C**. Scatter graph of RT-QulC ThT fluorescence intensity of αSyn^D^ seeding activity of saliva samples from PD patients and HC in the ss-subset. Scatter graph was plotted based on αSyn^D^ seeding activities in saliva samples from the PD patients and HC of the ss-subset shown in panel A. ThT fluorescence cutoff: 62,613. **** *p*<0.0001. **D**. ROC curve and AUC for saliva αSyn^D^ seeding activity comparisons between the PD patients and HC in a ss-subset. ROC curve and AUC value were obtained based on αSyn^D^ seeding activities in saliva of the PD patients and HC of the ss-subset shown in panel A. **E**. 3-D Plot toiIdentify optimal cutoff values for serum and saliva for maximum diagnostic accuracy for PD in the ss-subset shown in A. PD diagnostic accuracy was plotted against the RT-QuIC ThT fluorescence cutoff values of both serum and saliva in a 3-D plot, which identified the optimal αSyn^D^ seeding activity (ThT fluorescence) cutoff settings to achieve maximal diagnostic accuracy for PD as 52,960 for serum and 66,800 for saliva. The accuracy values were calculated by varying the cutting off values for both serum and saliva with the definition that a patient was considered positive for PD only when the αSyn^D^ seeding activities in both serum and saliva samples exceeded their respective cutoff values. **F**. ROC curve and AUC for PD diagnosis based on αSyn^D^ seeding activities in both serum and saliva of PD patients and HC in the ss-subset. ROC curve and AUC were obtained based on calculated sensitivity and specificity values when varying the ThT fluorescence cutoff values for both serum and saliva. The sensitivity and specificity values were calculated based on the same definition of PD positivity as described in panel E. R analysis of the paired serum and saliva αSyn^D^ seeding activity data of the ss-subset was in agreement with the ROC analysis. **** *p*<0.0001. SE, standard error. 95% CI, 95% confidence interval.

In contrast, when the aSyn-SAA data from both serum and saliva samples of the same patients were analyzed together, a much better diagnostic performance for all PD cases was achieved. For this analysis, a patient was defined as positive for PD if at least one of the samples (serum or saliva) was positive in the aSyn-SAA assay and a patient as negative for PD when both samples were negative in the aSyn-SAA assay. To identify the optimal ThT fluorescence cutoff values for the respective sample type (serum or saliva), a 3-D plot was made with values of accuracy, serum cutoff, and saliva cutoff (Figure 4E), which revealed relative ThT fluorescence units of 52,960 and 66,800 as the optimal cutoff values for serum and saliva samples, respectively. At these optimal cutoff values, a sensitivity of 95.83%, a specificity of 96.15%, and an accuracy of 93.75% were achieved for the diagnosis of all PD patients. If the specificity is set at 100%, a sensitivity of 91.67% and an accuracy of 91.67% are still attained. By varying the cutoff values for both serum and saliva samples and calculating the corresponding sensitivity and specificity values using the above definition for subjects’ PD positivity, we were able to generate a ROC curve for the combined serum-saliva aSyn^D^ seeding activity data, which showed an accuracy of 0.98 (AUC of ROC, 95% CI, 0.96-1.0, p<0.001) (Figure 4F).

Taken together, these results demonstrate that the diagnostic accuracy for PD based on both serum αSyn-SAA data and saliva αSyn-SAA data together is much better than using αSyn-SAA data from either sample type alone (Table 2).

### Clinical correlation of αSyn^D^ seeding activities in serum or saliva samples

We conducted extensive clinical assessments, including demographics (age, age at diagnosis, disease duration, gender), family history, mH&Y staging, MoCA, Schwab & England scoring, MDS-UPDRS assessments (parts 1, 2, 3, 4 and total score), PDQ39 (total, mobility, ADL, and cognitive impairment), ESS, HAM-A, HAM-D, orthostatic hypotension (self-reported and by vitals), as well as self-reported dementia, hyposmia (loss of smell), constipation, and RBD status.

We examined possible correlations between the aSyn-SAA status in serum and/or saliva samples of PD patients with the above clinical features and demographic factors (Table S1). When comparing the serum aSyn-SAA positivity between PD patients and HC subjects, significant differences were found for Schwab & England scale (*p*<0.001), PDQ39 scores and sub-scores [total (*p*=0.001), mobility (*p*=0.041), ADL (*p*<0.001), and cognitive impairment (*p*=0.06)], HAM-D (*p*=0.025), and patient self-reported hyposmia (loss of smell) (*p*=0.001) and constipation (*p*<0.001) (Table S1). For saliva aSyn-SAA positivity in PD and HC subjects, the *p*-value findings were similar, except that significant difference was also found for age (*p*=0.003), and the *p*-values were slightly higher for PDQ39-mobility (0.11 vs. 0.041) and HAM_D (0.078 vs. 0.025) (Table S1). A subset of the saliva donors (34 PD and 22 HC of cohort 2) was only included in the analysis for age, age at diagnosis, disease duration, sex, mH&Y, MoCA, and MSD-UPDRS Part 3 because other clinical information was not available for these subjects (Table S1).

Potential clinical correlations with aSyn^D^ seeding activities in serum or saliva samples were also examined by Pearson’s correlation analysis. αSyn^D^ seeding activities in the serum of PD patients were correlated significantly with MoCA (*p*=0.04, inversely) and HAM-D (*p*=0.03, positively), and weakly with PDQ39 cognitive impairment (*p*=0.07, positively) (Figure 5). No significant correlation was found with hyposmia (*p*=0.11), RBD (*p*=0.21), mH&Y (*p*=0.69), constipation (*p*=0.98), or any other features examined (Table S2). αSyn^D^ seeding activities in the saliva of PD patients were correlated significantly with age at diagnosis (*p*=0.023, inversely) and RBD status (*p*=0.04, inversely) (Figure 6). No significant correlation was found between αSyn^D^ seeding activities in the saliva of PD patients with MoCA (*p*=0.35), mH&Y (*p*=0.70), hyposmia (loss of smell, *p*=0.63), constipation (*p*=0.50) or any other features examined (Table S2).

**Figure 5.**
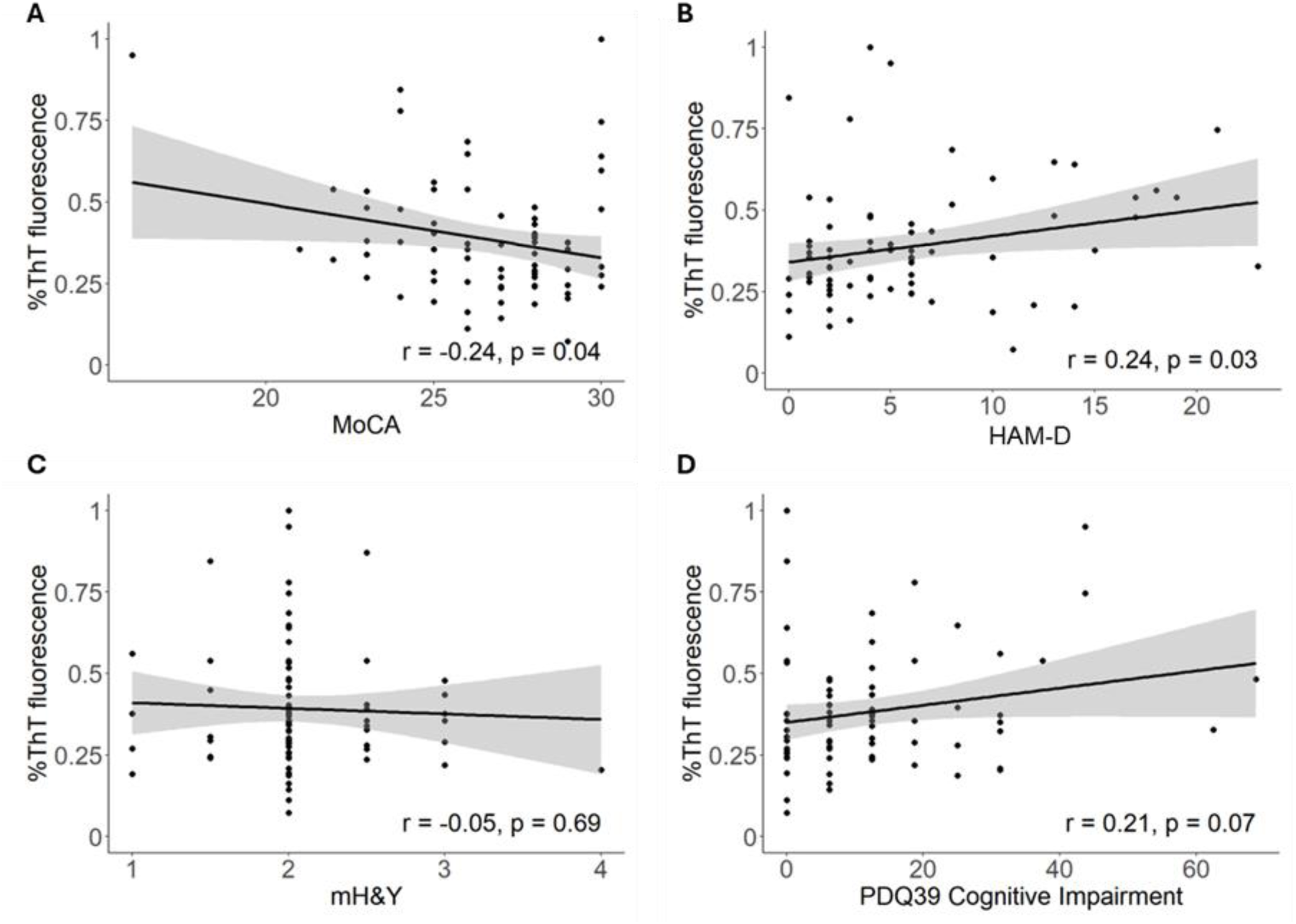
Serum αSyn^D^ Seeding Activities Correlate with MoCA, HAMD and PDQ39-Cognitive Impairment Scores among PD Patients. αSyn^D^ seeding activities (ThT fluorescence as a percentage of the maximum reading) in serum samples correlate inversely with MoCA score (**A**), positively with HAM-D score (**B**), and weakly positively with PDQ39-cognitive impairment score (**C**), but not with modified Hoehn & Yahr (mH&Y) (**D**) of PD patients. Linear regression lines with 95% confidence interval (gray shade) are shown.

**Figure 6.**
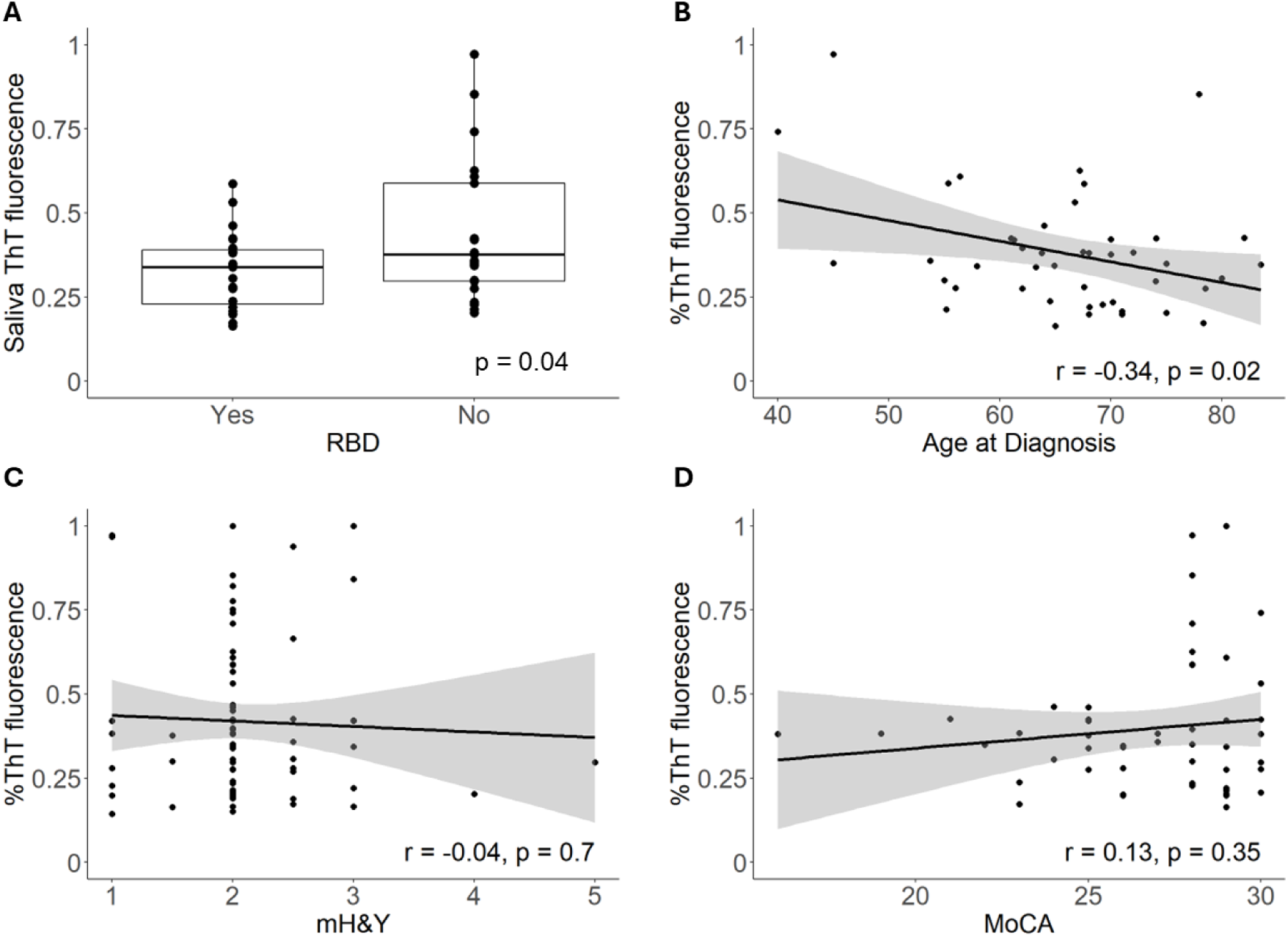
Saliva αSyn^D^ Seeding Activities Correlate with Age at Diagnosis and RBD among PD Patients. αSyn^D^ seeding activities (ThT fluorescence as a percentage of the maximum reading) in saliva samples correlate inversely with RBD status (**A**) and age at diagnosis (**B**), but not with modified Hoehn & Yahr (mH&Y) (**C**) or MoCA (**D**) of PD patients. Linear regression lines with 95% confidence interval (gray shade) are shown for **B-D**.

Subgroup analyses by sex (male or female) or age (<70 year or ≥70 year) were also performed (Tables S3 and S4).

For sex subgroup analyses of serum samples of PD patients, serum αSyn^D^ seeding activities were correlated inversely with MoCA in males (*p*=0.01) but not in females (*p*=0.70), weakly positively with PDQ39-cognitive impairment in females (*p*=0.07) but not in males (*p*=0.27), and positively with HAM-D score in females (*p*=0.04) but not in males (*p*=0.36) (Table S3). For age subgroup analyses of serum samples of PD patients, serum αSyn^D^ seeding activities were correlated weakly positively with MoCA in the ≥70 age group (*p*=0.07) but not in the <70 age group (*p*=0.15), positively with HAM-A in the <70 age group (*p*=0.04) but not in the ≥70 age group (*p*=0.40), positively with HAM-D in the <70 age group (*p*=0.01) but not in the ≥70 age group (*p*=0.92), positively with orthostatic hypotension by vitals in the <70 age group (*p*=0.01) but not in the ≥70 age group (*p*=0.31), inversely with ESS in the ≥70 age group (*p*=0.03) but not in the <70 age group (*p*=0.92), and inversely with RBD status in the ≥70 age group (*p*=0.03) but not in the <70 age group (*p*=0.56) (Table S3).

For sex subgroup analyses of saliva samples of PD patients, saliva αSyn^D^ seeding activities were correlated inversely with age at diagnosis in males (*p*=0.044) but not in females (*p*=0.13) (Table S4). For age subgroup analyses of saliva samples of PD patients, αSyn^D^ seeding activities were correlated inversely with age at diagnosis in the <70 age group (*p*=0.01) but not in the ≥70 age group (*p*=0.72), and inversely with Schwab & England in the ≥70 age group (p=0.02) but not in the <70 age group (*p*=0.11) (Table S4).

## DISCUSSION

Saliva and blood are distinct bodily fluids that have very different compositions and origins, and both have been shown to be good sources of αSyn^D^ seeding activities. While serum αSyn-SAA has been reported to show great performance in PD diagnosis that is comparable to (Okuzumi et al. 2023) or better than (Schaffer et al. 2024) the gold-standard CSF αSyn-SAA (Siderowf et al. 2023), these findings need to be reproduced and verified by other groups. We were unable to reproduce the performance of one of the prior serum αSyn-SAA reports (Okuzumi et al. 2023).

Saliva αSyn-SAA has also shown good sensitivity in detection of PD (76.0%-83.78%) and good specificity for healthy controls (82.61-94.4%) (Luan et al. 2022, Vivacqua et al. 2023), which are in line with our saliva αSyn-SAA data in this study (74.70% sensitivity and 97.92% specificity), confirming that saliva αSyn-SAA assay can detect αSyn^D^ seeding activities in saliva very well, but it may not be adequate as a stand-alone test for PD diagnosis.

We reasoned that PD patients who are αSyn-SAA negative in blood may be αSyn-SAA positive in their saliva and vice versa. To test this hypothesis, we examined paired serum and saliva samples from the ss-subset (48 PD and 26 HC) and demonstrated 0.98 in accuracy when the αSyn^D^ seeding activities in both serum and saliva were considered together, which is much better than using αSyn^D^ seeding activities in either serum or saliva alone (Figure 4 and Table 2).

This exciting finding indicates that, after further validations with larger sets of paired serum and saliva samples from more balanced cohorts of PD and HC subjects, αSyn^D^ seeding activities in serum and saliva together can be used as a valuable biomarker for highly sensitive, accurate, and minimally invasive PD diagnosis that can be more routinely implemented in the clinic and in clinical studies for PD diagnosis (Cardoso et al. 2023).

It is worth noting that the high diagnostic accuracy with αSyn-SAA data from both serum and saliva samples together was achieved only when the cutoff values for both sample types were set at high levels for high specificity in each sample type, suggesting that PD patients with low αSyn^D^ seeding activities in serum tend to have higher and detectable αSyn^D^ seeding activities in their saliva and vice versa. This finding suggests the same approach may also significantly improve the diagnostic accuracy for other synucleinopathies.

Our analyses also revealed significant correlations between αSyn^D^ seeding activities in serum or saliva and some clinical characteristics. When all serum samples or all saliva samples were analyzed together, αSyn^D^ seeding activities in serum of PD patients were correlated with MoCA (inversely) and HAM-D (positively), and weakly with PDQ-39 cognitive impairment (positively) (Figure 5), whereas αSyn^D^ seeding activities in saliva were correlated inversely with age at diagnosis and RBD status (Figure 6).

Age and sex subgroup analyses gave similar results but revealed age and sex dependence.

Notably, the serum αSyn^D^ seeding activities were correlated inversely with MoCA only with males and with the ≥70-year age group, positively correlated with HAM-D only with females and with the <70-year age group, and weakly positively corrected with PDQ39-cognitive impairment only with females (Table S3). Correlations with serum αSyn^D^ seeding activities found only in the subgroup analysis include with RBD status of the ≥70-year age group (inverse), with ESS score of the ≥70-year age group (inverse), with HAM-A of the <70-year age group (positive), with RBD status of the ≥70-year age group (inverse), and with orthostatic hypotension by vitals of the <70-year age group (positive) (Table S3). The saliva αSyn^D^ seeding activities were correlated inversely with age at diagnosis only with males and with the ≥70-year age group only with males and the <70-year age group, and inversely with Schwab & England scale that was not detected in the overall analysis (Table S4).

There have been other reports on correlations of αSyn^D^ seeding activities in various sample types with certain clinical features, but such correlations are often not corroborated by other studies, including this study. For example, the lack of correlation in with disease duration or motor stage (mH&Y) in this study contrasts with some other reports, including a recent report showing a one-way gradual decrease of serum αSyn^D^ seeding activity over time in a longitudinal study of 11 PD patients (Schaffer et al. 2024). CSF αSyn^D^ seeding activity was reported to corelate positively with olfactory deficit (Siderowf et al. 2023) that is common in PD patients (Haehner et al. 2011), and positively with UPDRS motor score and H&Y stage (Majbour et al. 2022). But other studies had different findings. For example, Orru et al (2021) detected an inverse correlation between CSF αSyn^D^ seeding activities and UPDRS motor score, directly opposite to the report by Majbour et al. (2022). Oftedal et al. (2023) also found no correlation between αSyn^D^ seeding activities and UPDRS 3 score and H&Y score in newly diagnosed PD patients. In another report (Kuzkina et al. 2021), skin αSyn-SAA data in PD patients showed positive correlation with disease duration, H&Y stage, RBD status, constipation, MCI, and non-motor symptoms score (NMSS) and inverse correlation with MoCA score. Saliva αSyn^D^ seeding activities were also reported to be positively correlated with MDS-UPDRS 3 score and NMSS score (Vivacqua et al. 2023), but our saliva αSyn-SAA data failed to show significant correlation with MDS-UPDRS 3 or several non-motor assessment scores, such as MoCA and PDQ-39 total. The reasons for the differences amongst studies in clinical correlations with αSyn^D^ seeding activity are not clear, but might reflect differences in study populations, sample types used, and αSyn-SAA parameters.

There have also been discussions of using CSF αSyn-SAA for staging of PD (Cardoso et al. 2023). Patients are often resistant to lumbar puncture for diagnosis of prodromal or early-stage PD since it may not change clinical management and the diagnosis should reveal itself with time. However, a prompt and accurate diagnosis and staging of early and prodromal PD is important for enrollment in neuroprotective studies. Our data suggest that serum/saliva αSyn-SAA is also valuable for PD diagnosis. If it proves to be sensitive and specific for early/prodromal PD, it could serve as a patient-friendly surrogate of CSF αSyn-SAA for early pathological diagnosis of PD.

## LIMITATIONS

This study has some limitations. First, the number of serum and saliva samples from PD and healthy subjects examined, especially those from healthy subjects, are still relatively small and from one cohort of PD and HC subjects only. Second, the paired serum-saliva samples from the same subjects are relatively small in total number. Third, the HC subjects were primarily females while the PD patients had slightly more males than females. Fourth, the impact of LRRK2 genotype status and UPSIT smell test results on αSyn-SAA outcome in the PPMI CSF cohort study reported by Siderowf et al. (2023) were not assessed in this study. Fifth, no serum or saliva samples from non-PD synucleinopathy patients were included. Our findings need to be validated with a larger number of paired serum-saliva samples from gender-balanced cohorts of PD and HC subjects from a variety of racial and socioeconomic backgrounds and encompassing all clinical stages, as well as large balanced cohorts of non-PD synucleinopathy patients (such as MSA, DLB and PDP) and prodromal patients (such as RBD).

## CONCLUSIONS

Our study suggests that RT-QuIC analysis of αSyn^D^ seeding activities in both serum and saliva samples together can be used as a minimally invasive and reliable biomarker for highly sensitive and accurate PD diagnosis in routine clinical practice and clinical studies, and that αSyn^D^ seeding activities in serum and saliva samples are differentially correlated with various clinical characteristics in an age and sex-dependent manner.

## Data Availability

All data produced in the present work are contained in the manuscript.

## Author Contributions

Drs. Kong, Wang, and Gunzler had full access to all of the data in the study and take responsibility for the integrity of the data and the accuracy of the data analysis. Drs. Kong, Wang, Gunzler and Chen are co-senior authors. Study concept and design: Kong, Wang, and Chen. Acquisition, analysis, or interpretation of data: All authors. Drafting of the manuscript: Kong, Wang, Gunzler, and Chen. Critical revision of the manuscript for important intellectual content: All authors. Statistical analysis: H.J.K & Drs. Bebek, Wang, and Kong. Obtained funding: Kong, Wang, Gunzler, and Chen. Administrative, technical, or material support: All authors. Study supervision: Kong, Wang, Gunzler, and Chen.

