## Supplementary material for "A minimally Invasive Biomarker for Sensitive and Accurate Diagnosis of Parkinson’s Disease": Kong Manuscript Supplemental Data.pdf

### Supplementary Data

#### Supplementary Tables

**Table S1.** Clinical characteristics of all PD and HC subjects by αSyn-SAA status in serum or saliva

| | **Parkinson's disease (PD)** | | | | | | **Healthy controls (HC)** | | | |
|---|---|---|---|---|---|---|---|---|---|---|
| | **Serum** | | | **Saliva** | | | **Serum** | | **Saliva** | |
| | SAA+ | All PD | *p* value | SAA+ | All PD | *p* value | SAA+ | All HC | SAA+ | All HC |
| | n=66 | n=82 | All PD vs HC | n=40 (62*) | n=49 (83*) | All PD vs HC | n=4 | n=42 | n=1 (1*) | n=26 (48*) |
| Age (years), mean (range) | 69.35 (44-88) | 69.21 (44-88) | 0.114 | 69.29 (49-85) | 69.58 (49-87) | **0.003** | 69 (59-73) | 66.55 (44-81) | 66 (66-66) | 64.71 (30-81) |
| Age at diagnosis (years), mean (range) | 66.31 (39.00-88.42) | 65.84 (39.00-88.42) | 0.733 | 65.15 (40.0-83.5) | 65.86 (40.00-83.50) | 0.556 | NA | 63.86 (51.67-74.00) | NA | 69.96 (65.92-74.00) |
| Disease duration, years (range) | 4.96 (0-17) | 5.05 (0-17) | NA | 6.76 (0-31) | 6.18 (0-31) | NA | NA | NA | NA | NA |
| Sex | | | **0.004** | | | **0.006** | | | | |
| Male, number (%) | 35 (53.0) | 45 (54.9) | | 31 (50.0) | 46 (55.4) | | 0 (0.0) | 11 (26.2) | 0 (0.0) | 14 (29.2) |
| Female, number (%) | 31 (47.0) | 37 (45.1) | | 31 (50.0) | 37 (44.6) | | 4 (100.0) | 31 (73.8) | 1 (100.0) | 34 (70.8) |
| mH&Y, mean (SD) | 2.05 (0.44) | 2.05 (0.48) | NA | 2.13 (0.59) | 2.09 (0.61) | NA | NA | NA | NA | NA |
| Schwab & England, mean (SD) | 90.23 (7.46) | 90.55 (7.11) | **<0.001** | 92.24 (6.85) | 91.12 (8.12) | **<0.001** | 97.5 (5.0) | 98.97 (3.07) | 100 (0.0) | 99.17 (2.82) |
| MoCA, mean (SD) | 26.49 (2.79) | 26.69 (2.60) | 0.989 | 26.69 (3.20) | 26.74 (3.04) | 0.426 | 27.5 (1.29) | 26.70 (2.28) | 20 (0.0) | 27.24 (2.40) |
| MDS-UPDRS | | | | | | | | | | |
| Part 1, mean (SD) | 7.69 (5.79) | 7.34 (5.82) | NA | 7.35 (6.25) | 7.58 (6.42) | NA | NA | NA | NA | NA |
| Part 2, mean (SD) | 8.98 (5.65) | 8.69 (5.75) | NA | 8.26 (6.15) | 8.16 (5.91) | NA | NA | NA | NA | NA |
| Part 3, mean (SD) | 31.78 (9.58) | 31.46 (10.62) | NA | 28.96 (11.32) | 29.00 (11.27) | NA | NA | NA | NA | NA |
| Parts 123, mean (SD) | 48.67 (16.75) | 47.64 (17.85) | NA | 45.64 (17.92) | 45.59 (18.07) | NA | NA | NA | NA | NA |
| Part 4 mean (SD) | 1.80 (2.46) | 1.81 (2.47) | NA | 1.88 (2.57) | 1.62 (2.49) | NA | NA | NA | NA | NA |
| Total score | 50.50 (17.54) | 49.47 (18.76) | NA | 47.58 (18.64) | 47.25 (19.03) | NA | NA | NA | NA | NA |
| PDQ39 | | | | | | | | | | |
| total, mean (SD) | 95.99 (84.03) | 94.84 (82.14) | **0.001** | 79.88 (71.46) | 87.17 (82.15) | **0.003** | 2.78 (4.81) | 35.53 (65.15) | 8.3 (0.0) | 23.89 (31.10) |
| mobility, mean (SD) | 9.42 (11.81) | 9.05 (11.82) | **0.041** | 6.76 (10.33) | 7.50 (10.98) | 0.110 | 6.25 (12.5) | 4.21 (10.64) | 0.0 (0.0) | 3.21 (7.29) |
| ADL, mean (SD) | 14.79 (13.32) | 14.47 (14.15) | **<0.001** | 11.76 (11.94) | 12.97 (13.60) | **<0.001** | 0.0 (0.0) | 1.79 (4.77) | 0.0 (0.0) | 1.39 (4.03) |
| cognitive impairment, mean (SD) | 13.33 (14.98) | 12.75 (14.20) | **0.006** | 12.32 (14.96) | 13.64 (16.52) | 0.069 | 3.12 (6.25) | 5.54 (8.13) | 0.0 (0.0) | 6.55 (8.50) |
| Patient self-report | | | | | | | | | | |
| Dementia, number (%) | 10 (15.2) | 10 (12.2) | 0.395 | 11 (17.7) | 12 (14.5) | 0.065 | 1 (25.0) | 2 (5.3) | 0 (0.0) | 1 (2.3) |
| Hyposmia, number (%) | 33 (50.0) | 37 (45.1) | **<0.001** | 17 (44.7) | 21 (42.9) | **0.001** | 0 (0.0) | 1 (2.6) | 0 (0.0) | 0 (0.0) |
| Constipation, number (%) | 31 (47.7) | 38 (46.9) | **<0.001** | 17 (44.7) | 24 (49.0) | **0.001** | 0 (0.0) | 2 (5.3) | 0 (0.0) | 1 (4.5) |
| ESS, mean (SD) | 5.97 (3.71) | 5.95 (3.68) | 0.119 | 6.24 (3.72) | 6.66 (3.80) | 0.075 | 3.50 (2.38) | 4.87 (2.98) | 2.00 (0.0) | 5.04 (3.13) |
| HAM-A, Mean (SD) | 7.89 (7.23) | 7.70 (7.06) | 0.162 | 7.63 (7.55) | 8.00 (7.69) | 0.387 | 8.25 (13.87) | 5.85 (5.97) | 3.00 (0.0) | 6.40 (6.73) |
| HAM-D, mean (SD) | 5.97 (5.64) | 5.75 (5.44) | **0.025** | 5.64 (5.86) | 6.00 (5.83) | 0.078 | 4.25 (7.23) | 3.51 (3.98) | 2 (0.0) | 3.60 (4.31) |
| RBD, number (%) | 29 (48.3) | 35 (46.1) | **<0.001** | 17 (50.0) | 23 (52.3) | **<0.001** | 0 (0.0) | 0 (0.0) | 0 (0.0) | 0 (0.0) |

*Total number of saliva donors that include a subset of subjects with information on age, age at diagnosis, disease duration, sex, mH&Y, MoCA, and MSD-UPDRS 3 only. RT-QuIC ThT fluorescence cutoff: 52,105 for serum and 62,163 for saliva. SAA +: patients with positive αSyn-SAA in the indicated sample.

**Table S2**. Clinical correlation analysis by αSyn$^{D}$ seeding activities in serum or saliva of PD patients

| | **Serum** (n = 82) | | **Saliva** (n = 83) | |
|---|---|---|---|---|
| **Pearson's correlation** | ***r*** | ***p*** | ***r*** | ***p*** |
| Age | -0.12 | 0.28 | -0.10 | 0.38 |
| Disease duration | 0.05 | 0.63 | 0.03 | 0.78 |
| Age at diagnosis | -0.15 | 0.20 | **-0.34** | **0.02** |
| Schwab & England | -0.14 | 0.21 | 0.025 | 0.87 |
| mH&Y | -0.05 | 0.69 | -0.04 | 0.70 |
| MoCA | **-0.24** | **0.04** | 0.13 | 0.35 |
| MDS-UPDRS | | | | |
| Part 1 | 0.05 | 0.65 | -0.07 | 0.64 |
| Part 2 | 0.01 | 0.93 | -0.07 | 0.66 |
| Part 3 | 0.02 | 0.85 | -0.05 | 0.67 |
| Part 123 | 0.03 | 0.81 | -0.12 | 0.43 |
| Part 4 | 0.04 | 0.73 | -0.05 | 0.76 |
| Total | 0.03 | 0.79 | -0.12 | 0.43 |
| PDQ39 | | | | |
| Total | 0.08 | 0.52 | -0.07 | 0.68 |
| Mobility | 0.01 | 0.93 | -0.15 | 0.34 |
| ADL | <0.01 | 0.97 | -0.17 | 0.28 |
| Cognitive impairment | **0.21** | **0.07** | -0.07 | 0.66 |
| ESS | -0.15 | 0.19 | -0.23 | 0.13 |
| HAM-A | 0.13 | 0.25 | 0.05 | 0.76 |
| HAM-D | **0.24** | **0.03** | 0.05 | 0.75 |
| **ANOVA** | **Higher mean** | ***p*** | **Higher mean** | ***p*** |
| Sex | female | 0.95 | Female | 0.31 |
| RBD | no | 0.21 | **no** | **0.04** |
| Orthostatic hypotension (vitals) | yes | 0.72 | no | 0.84 |
| Patient self-report | | | | |
| Hyposmia | yes | 0.11 | no | 0.63 |
| Orthostatic hypotension | yes | 0.97 | yes | 0.66 |
| Constipation | yes | 0.98 | Yes | 0.50 |
| Dementia | yes | 0.16 | Yes | 0.27 |

**Table S3**. Correlation subgroup analysis by sex or age by serum αSyn$^D$ seeding activities of PD patients

| | **Sex** | | | | **Age** | | | |
|---|---|---|---|---|---|---|---|---|
| | **Female** (n = 37) | | **Male** (n = 46) | | **< 70 years** (n = 39) | | **≥ 70 years** (n = 44) | |
| **Pearson's correlation** | *r* | *p* | *r* | *p* | *r* | *p* | *r* | *p* |
| Age | -0.17 | 0.33 | -0.09 | 0.57 | 0.03 | 0.84 | -0.16 | 0.30 |
| Disease duration | -0.03 | 0.88 | 0.12 | 0.45 | 0.01 | 0.95 | 0.11 | 0.49 |
| Age at diagnosis | -0.18 | 0.30 | -0.12 | 0.44 | 0.01 | 0.96 | -0.14 | 0.39 |
| Schwab & England | -0.18 | 0.28 | -0.10 | 0.51 | -0.21 | 0.21 | -0.12 | 0.43 |
| mH&Y | -0.02 | 0.93 | -0.07 | 0.66 | 0.05 | 0.75 | -0.08 | 0.63 |
| MoCA | -0.07 | 0.70 | **-0.38** | **0.01** | -0.25 | 0.15 | **-0.29** | **0.07** |
| MDS-UPDRS | | | | | | | | |
| Part 1 | 0.04 | 0.82 | 0.07 | 0.68 | 0.27 | 0.12 | -0.21 | 0.19 |
| Part 2 | 0.09 | 0.62 | -0.02 | 0.89 | 0.14 | 0.44 | -0.04 | 0.78 |
| Part 3 | 0.25 | 0.16 | -0.15 | 0.35 | 0.24 | 0.16 | -0.15 | 0.35 |
| Part 123 | 0.19 | 0.30 | -0.07 | 0.67 | 0.29 | 0.09 | -0.12 | 0.45 |
| Part 4 | 0.09 | 0.62 | <-0.01 | 0.98 | 0.08 | 0.63 | 0.03 | 0.85 |
| Total | 0.19 | 0.30 | -0.07 | 0.68 | 0.29 | 0.09 | -0.14 | 0.39 |
| PDQ39 | | | | | | | | |
| Total | 0.10 | 0.62 | 0.09 | 0.62 | 0.32 | 0.09 | -0.14 | 0.43 |
| Mobility | 0.09 | 0.63 | -0.08 | 0.64 | 0.22 | 0.21 | -0.03 | 0.84 |
| ADL | 0.07 | 0.69 | -0.03 | 0.86 | 0.17 | 0.34 | -0.10 | 0.55 |
| Cognitive impairment | **0.32** | **0.07** | 0.17 | 0.27 | 0.23 | 0.18 | 0.16 | 0.33 |
| ESS | -0.20 | 0.26 | -0.11 | 0.50 | 0.02 | 0.92 | **-0.33** | **0.03** |
| HAM-A | 0.21 | 0.23 | 0.06 | 0.69 | **0.36** | **0.04** | -0.13 | 0.40 |
| HAM-D | **0.36** | **0.04** | 0.14 | 0.36 | **0.43** | **0.01** | 0.02 | 0.92 |
| **ANOVA** | **Higher mean** | ***p*** | **Higher mean** | ***p*** | **Higher mean** | ***p*** | **Higher mean** | ***p*** |
| Sex | - | - | - | - | male | 0.56 | female | 0.61 |
| RBD | No | 0.17 | yes | 0.68 | yes | 0.56 | **no** | **0.03** |
| Orthostatic hypotension (vitals) | yes | 0.40 | no | 0.78 | **yes** | **0.01** | no | 0.31 |
| Patient self-report | | | | | | | | |
| Hyposmia | yes | 0.67 | yes | 0.09 | yes | 0.09 | yes | 0.41 |
| Orthostatic hypotension | no | 0.48 | yes | 0.71 | yes | 0.76 | yes | 0.83 |
| Constipation | no | 0.76 | yes | 0.78 | yes | 0.35 | no | 0.63 |
| Dementia | yes | 0.24 | yes | 0.33 | yes | 0.26 | yes | 0.28 |

**Table S4**. Correlation subgroup analysis by sex or age by saliva αSyn$^D$ seeding activities of PD patients

| | **Sex** | | | | **Age** | | | |
|---|---|---|---|---|---|---|---|---|
| | **Female** (n = 37) | | **Male** (n = 46) | | **< 70 years** (n = 39) | | **≥ 70 years** (n = 44) | |
| **Pearson's correlation** | *r* | *p* | *r* | *p* | *r* | *p* | *r* | *p* |
| Age | <0.01 | 0.99 | -0.19 | 0.21 | -0.13 | 0.44 | -0.17 | 0.28 |
| Disease duration | 0.07 | 0.67 | -0.01 | 0.93 | -0.11 | 0.50 | 0.12 | 0.43 |
| Age at diagnosis | -0.33 | 0.13 | **-0.42** | **0.04** | **-0.52** | **0.01** | 0.08 | 0.72 |
| Schwab & England | -0.08 | 0.70 | 0.13 | 0.53 | 0.33 | 0.11 | **-0.47** | **0.02** |
| mH&Y | 0.25 | 0.15 | -0.17 | 0.27 | -0.10 | 0.56 | <-0.01 | 0.97 |
| MoCA | 0.07 | 0.77 | 0.15 | 0.46 | 0.18 | 0.37 | 0.08 | 0.72 |
| MDS-UPDRS | | | | | | | | |
| Part 1 | 0.17 | 0.45 | -0.22 | 0.31 | -0.17 | 0.45 | 0.11 | 0.64 |
| Part 2 | 0.08 | 0.73 | -0.13 | 0.56 | -0.16 | 0.46 | 0.03 | 0.89 |
| Part 3 | 0.12 | 0.50 | -0.13 | 0.39 | -0.02 | 0.89 | -0.05 | 0.75 |
| Part 123 | 0.16 | 0.49 | -0.32 | 0.13 | -0.18 | 0.42 | -0.02 | 0.95 |
| Part 4 | 0.06 | 0.79 | -0.16 | 0.47 | -0.08 | 0.73 | <0.01 | 0.99 |
| Total | 0.16 | 0.50 | -0.33 | 0.12 | -0.18 | 0.40 | -0.01 | 0.96 |
| PDQ39 | | | | | | | | |
| Total | 0.33 | 0.18 | -0.28 | 0.21 | -0.22 | 0.32 | 0.18 | 0.47 |
| Mobility | -0.12 | 0.60 | -0.18 | 0.42 | -0.23 | 0.30 | -0.08 | 0.73 |
| ADL | -0.10 | 0.68 | -0.20 | 0.35 | -0.19 | 0.40 | -0.18 | 0.50 |
| Cognitive impairment | 0.35 | 0.12 | -0.22 | 0.32 | -0.12 | 0.58 | -0.04 | 0.85 |
| ESS | -0.06 | 0.78 | -0.36 | 0.09 | -0.34 | 0.11 | -0.11 | 0.64 |
| HAM-A | 0.33 | 0.13 | -0.19 | 0.38 | -0.01 | 0.95 | 0.08 | 0.72 |
| HAM-D | 0.35 | 0.13 | -0.18 | 0.43 | 0.02 | 0.93 | <0.01 | 0.97 |
| **ANOVA** | **Higher mean** | ***p*** | **Higher mean** | ***p*** | **Higher mean** | ***p*** | **Higher mean** | ***p*** |
| Sex | - | - | - | - | female | 0.81 | female | 0.14 |
| RBD | no | 0.13 | no | 0.34 | no | 0.13 | no | 0.15 |
| Orthostatic hypotension (vitals) | no | 0.37 | yes | 0.25 | yes | 0.28 | no | 0.62 |
| Patient self-report | | | | | | | | |
| Hyposmia | no | 0.61 | no | 0.92 | no | 0.09 | yes | 0.36 |
| Orthostatic hypotension | no | 0.84 | no | 0.70 | no | 0.37 | yes | 0.79 |
| Constipation | yes | 0.68 | yes | 0.81 | yes | 0.67 | no | 0.69 |
| Dementia | yes | 0.98 | yes | 0.15 | yes | 0.37 | yes | 0.51 |

#### Supplementary Figures

**Figure S1.** Comparison of αSyn$^{D}$ Seeding Activity in All Serum or Saliva Samples from Patients with Probable or Possible PD and Healthy Control (HC) by RT-QuIC.

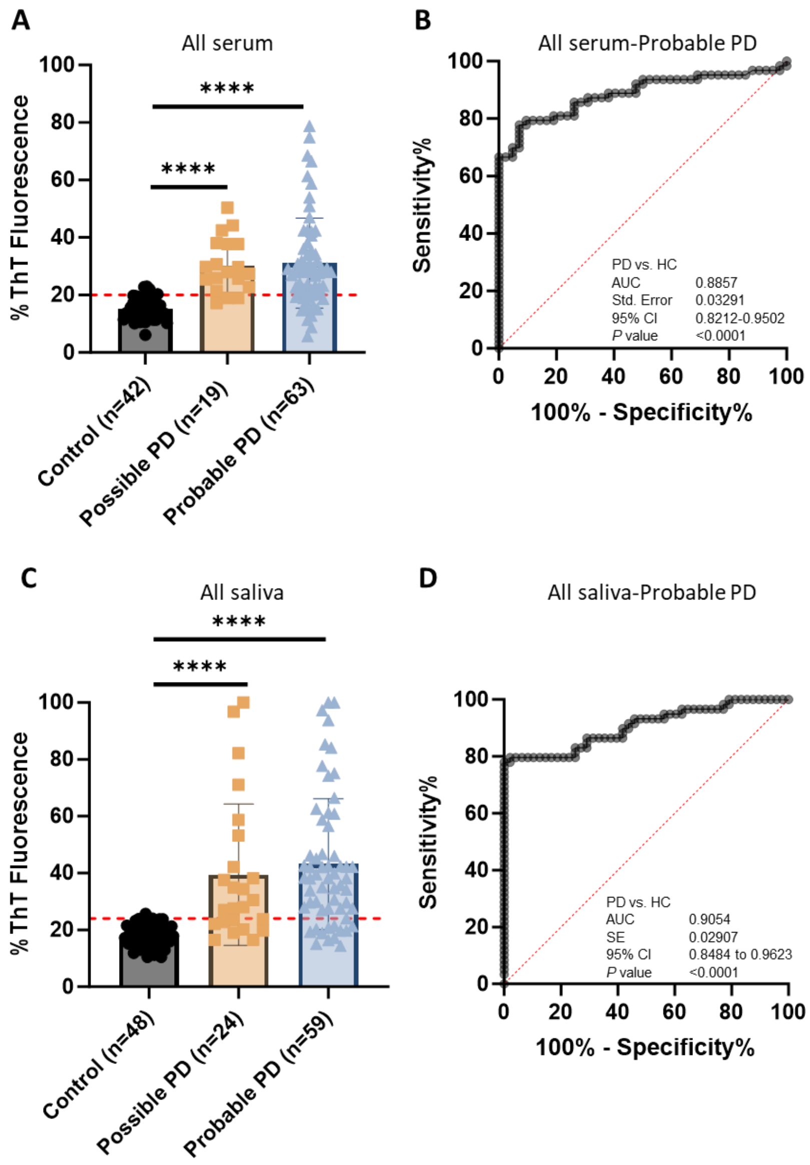

**A.** Scatter graph of RT-QuIC ThT fluorescence intensity of αSyn$^{D}$ seeding activity of serum samples from all PD patients and HC subjects. Scatter graph of the average of the peak thioflavin T (ThT) fluorescence in quadruplicate wells as a percentage of the maximum fluorescence (%ThT fluorescence) of serum samples from 19 possible PD patients, 63 probable PD patients, and 42 HC subjects by RT-QuIC assay. ThT fluorescence cutoff: 52,105. **** $p<0.0001$. **B.** ROC curve and AUC for αSyn$^{D}$ seeding activity comparisons between serum samples of the probable PD patients and HC subjects. ROC curve and AUC value for αSyn$^{D}$ seeding activities in the serum of all probable PD patients and HC subjects are shown. SE, standard error. 95% CI, 95% confidence interval. **C.** Scatter graph of RT-QuIC ThT fluorescence intensity of αSyn$^{D}$ seeding activities of saliva samples from all PD patients and HC subjects. Scatter graph of the average of the peak thioflavin T (ThT) fluorescence in quadruplicate wells as a percentage of the maximum possible fluorescence (%ThT fluorescence) of saliva samples from 24 possible PD patients, 59 probable PD patients, and 48 HC subjects by RT-QuIC assay. ThT fluorescence cutoff: 62,613. **** $p<0.0001$. **D.** ROC curve and AUC for αSyn$^{D}$ seeding activity comparisons between saliva samples of the 59 probable PD patients and 48 HC subjects. ROC curve and AUC value for αSyn$^{D}$ seeding activities in the serum of all probable PD patients and HC subjects are shown. SE, standard error. 95% CI, 95% confidence interval.

**Figure S2.** Comparison of αSyn$^D$ Seeding Activity in Serum or Saliva Samples from Patients with Probable or Possible PD and Healthy Control (HC) of the ss-subset by RT-QuIC.

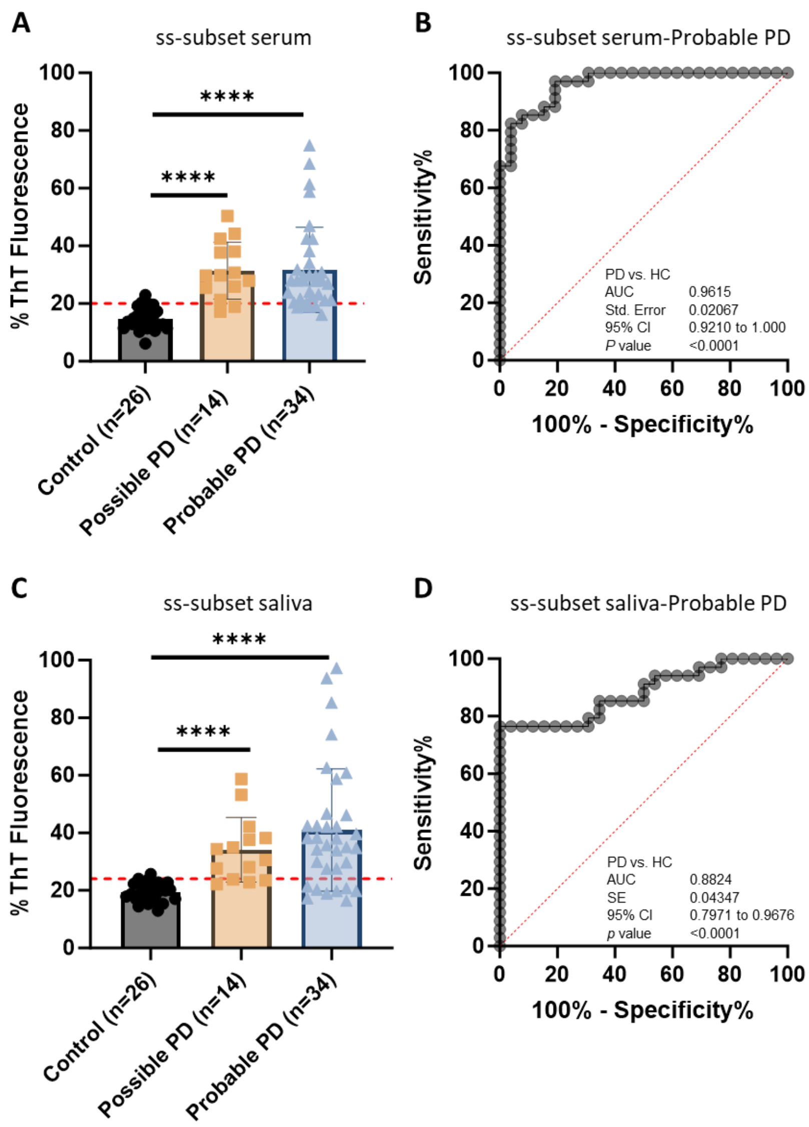

**A.** Scatter graph of RT-QuIC ThT fluorescence intensity of αSyn$^D$ seeding activity of serum samples from PD patients and HC subjects of the ss-subset. Scatter graph of the average of the peak thioflavin T (ThT) fluorescence in quadruplicate wells as a percentage of the maximum possible fluorescence (%ThT fluorescence) of serum samples from 14 possible PD patients, 34 probable PD patients, and 26 HC subjects by RT-QuIC assay. ThT fluorescence cutoff: 52,105. **** $p<0.0001$. **B.** ROC curve and AUC for αSyn$^D$ seeding activity comparisons between serum samples of probable PD patients and HC subjects of the ss-subset. ROC curve and AUC value for αSyn$^D$ seeding activities in the serum of probable PD patients and HC subjects of the ss-subset are shown. SE, standard error. 95% CI, 95% confidence interval. **C.** Scatter graph of RT-QuIC ThT fluorescence intensity of αSyn$^D$ seeding activities of saliva samples from PD patients and HC subjects of the ss-subset. Scatter graph of the average of the peak thioflavin T (ThT) fluorescence in quadruplicate wells as a percentage of the maximum fluorescence (%ThT fluorescence) of saliva samples from 14 possible PD patients, 34 probable PD patients, and 26 HC subjects by RT-QuIC assay. ThT fluorescence cutoff: 62,613. **** $p<0.0001$. **D.** ROC curve and AUC for αSyn$^D$ seeding activity comparisons between saliva samples of the 34 probable PD patients and 26 HC subjects. ROC curve and AUC value for αSyn$^D$ seeding activities in the serum of probable PD patients and HC subjects in the ss-subset are shown. SE, standard error. 95% CI, 95% confidence interval.
